# Racial and Ethnic Differences in Obesity Treatment in the Veterans Affairs Healthcare System

**DOI:** 10.1101/2024.02.02.24302244

**Authors:** Rebecca L. Tisdale, Tariku J. Beyene, Wilson Tang, Paul Heidenreich, Steven Asch, Celina M. Yong

**Affiliations:** Veterans Affairs Palo Alto Health Care System, Palo Alto, CA; Department of Medicine, Stanford University School of Medicine, Stanford, California

## Abstract

**Background:** The burden of obesity in the United States and in the Veterans Affairs (VA) population disproportionately affects minoritized individuals. Major advances in the treatment of obesity have emerged in the last decade, including incretin-based injectables like semaglutide, but it is not clear that they are being distributed equitably. We examined the role that race and ethnicity play in the receipt of a lifestyle change program, medications, and surgical treatment for obesity.

**Methods:** We analyzed data from patients with BMI ≥27 in the national VA Healthcare System from 2018-2022. We used multivariate logistic regression to evaluate the association between race/ethnicity (American Indian/Alaska Native [AI/AN], Asian/Native Hawaiian/Pacific Islander [NHOPI], Non-Hispanic Black, Hispanic/Latino, Non-Hispanic White) and use of weight loss interventions (VA lifestyle change program, medication prescriptions, bariatric surgery), adjusting for age, gender, comorbidities, and diagnosis year.

**Results:** Among Veterans with BMI ≥27 (n=2,423,070), 38.8% had Class 1 obesity (BMI 30-34.9), 18.1% had Class 2 obesity (BMI 35-39.9), and 9.8% had Class 3 obesity (BMI>40), with significant differences by race/ethnicity (p<0.01). Across the cohort, 7.7% participated in the lifestyle change program, 7.9% received weight loss medications, and 0.1% underwent bariatric surgery. Compared to Non-Hispanic White patients, Veterans belonging to many racial/ethnic minority groups were more likely to participate in the lifestyle change program (Asian/NHOPI Veterans, 1.12 [95% CI 1.06-1.19]; Non-Hispanic Black Veterans, adj OR 1.24 [95% confidence interval [CI] 1.22-1.26]; Hispanic/Latino Veterans, adj OR 1.17 [95% CI 1.14-1.20]) and less likely to receive weight loss medications (AI/AN Veterans, adj OR 0.84 [95% CI 0.77-0.92]; Asian/NHOPI Veterans: adj OR 0.94 [95% CI 0.89-0.999]; Non-Hispanic Black Veterans, adj OR 0.75 [95% CI 0.74-0.76]; Hispanic/Latino Veterans, adj OR 0.94 [95% CI 0.91-0.97]). Black Veterans were also less likely to undergo bariatric surgery (adj OR 0.79 [95% CI 0.69-0.89]).

**Conclusions:** Among Veterans with obesity, rates of treatment across all modalities are low. Inequities in treatment approach by race/ethnicity suggest areas for focused intervention to close gaps in care.

## Introduction

The past decade has seen major advances in the treatment of obesity, most notably the emergence of the highly effective incretin-based injectable medications semaglutide and tirzepatide (1,2). These advances have been motivated by the emergence of obesity as one of the most critical public health problems in the United States. With a prevalence of 42% of the overall population, obesity leads to over 300,000 premature deaths per year, of which more than two-thirds are caused by cardiovascular disease, and medical costs of $173 billion annually (3–5).

This burden is not distributed equally across the population. Historically marginalized groups suffer disproportionately from obesity, with higher-than-average prevalence among Black and Hispanic individuals in particular (3,6). Native American/Alaska Native and Native Hawaiian/Pacific Islander Veterans have also been shown to have disproportionately high prevalence of obesity (6). Increasing burden of disease coupled with disparities in prevalence threaten to widen health inequities in the United States over time (7), yet the extent to which these disparities are addressed by new treatments is uncertain.

Any improvement in these disparities in obesity itself depends on allocating obesity treatments fairly. Incretin-based injectable medications join the existing arsenal of evidence-based, guideline-recommended obesity treatments, including behavioral weight loss programs, older anti-obesity medications, and bariatric surgery (8–12), all of which were underutilized in the era prior to incretin-based therapy (13–19). As marginalized populations frequently receive healthcare innovations slower and later than more resource-rich counterparts (20,21)—and given inequities in use of existing obesity treatments (18,19,22–25)—it is far from certain that these next-generation medications are diffusing equitably. Indeed, early experiences suggest the opposite (26); however, inequalities are often attributed to insurance coverage or ability to pay.

The Veterans Health Administration (VA), with a high prevalence of obesity and diverse patient population (6), offers a unique opportunity to understand whether these inequalities persist within an integrated healthcare system where patients do not face insurance barriers to care. In this study, we describe the use of obesity treatments between 2018 and 2023 among VA patients with overweight or obesity, and differences therein by race and ethnicity.

## Methods

### Study Population

VA is the nation’s largest integrated healthcare system, caring for over 9 million Veterans across 172 VA medical centers and associated clinics across the United States (27). Using body mass index (BMI) data from VA’s Corporate Data Warehouse, which includes patient demographic data, diagnoses, procedures, and pharmacy dispensing records, we identified all VA patients aged ≥18 with two or more weight and height measurements at least 7 days apart and at least one BMI ≥27 from January 1, 2018 to December 31, 2022. We collected data for these patients from January 1, 2018 through December 31, 2023 (i.e., our observation period extended for one year beyond our cohort enrollment period). We excluded those for whom the initial BMI ≥27 was made outside of the United States and those with BMI ≥80 (likely due to entry error). Given that eligibility for most obesity treatments in the VA begins at a BMI of 27, we defined overweight as the group with BMI 27-29.9 (which is a subset of the full range of overweight defined by the CDC as BMI 25-29.9 [10]). Class 1 obesity was defined as BMI 30-34.9, class II obesity as BMI 35-39.9, and class III obesity as BMI ≥40. Race/ethnicity was obtained from the electronic health record (<5% missing in the national VA cohort), which categorizes patients into the following groups: American Indian/Alaska Native (AI/AN), Black Non-Hispanic, Asian, Hispanic or Latino, Native Hawaiian or Pacific Islander (NHOPI), and White Non-Hispanic. Patients with missing race/ethnicity were excluded. If patients had both Hispanic or Latino ethnicity and White or Black race, ethnicity took precedence for classification purposes. Due to low numbers of Asian and NHOPI patients, we grouped them into a single category (“Asian/NHOPI”) for our statistical models. Black Non-Hispanic and White Non-Hispanic Veterans will henceforth be referred to as Black and White Veterans for simplicity.

We extracted data on selected comorbidities based on *International Classification of Diseases, Tenth Revision* (ICD-10) diagnosis codes for the pertinent conditions in the two years prior to the study period (**Table S1**). The exception to this was diabetes status, given its relevance for medication prescriptions, which was defined as a diagnosis of diabetes in the two years prior to the study period or at any time during the study period.

### Interventions Studied

We included data on use of three categories of interventions for obesity: MOVE! (VA’s lifestyle counseling intervention), medications for obesity, and bariatric surgeries.

The MOVE! behavioral change program is offered in every VA medical center, and Veterans are not charged copayments to participate (29); it comprises a comprehensive lifestyle intervention with support and strategies to change eating patterns, physical activity, and related domains (30,31). We captured MOVE! visits via VA-specific “stop codes” corresponding to these visits. We considered Veterans to be MOVE! users if they attended one or more sessions, since this implied they had some degree of access to the program. We only counted new attendance by excluding anyone who had participated in the prior five years.

Medications included phentermine-topiramate, phentermine monotherapy, orlistat, bupropion-naltrexone, and semaglutide, which included both the Ozempic and Wegovy forms. Use of a medication was defined as having at least one pharmacy fill of a medication.

Bariatric surgical interventions included adjustable gastric band, sleeve gastrectomy, and roux-en-y gastric bypass; CPT and ICD10 codes corresponding to these surgeries are presented in **Table S2**. Analogous to the way we defined MOVE! visits, we counted surgeries for patients without a history of bariatric surgery within the last five years.

In addition to capturing use of each category of intervention alone, we collected data on use of combinations of interventions (e.g., MOVE! and bariatric surgery).

### Statistical Analysis

We describe the breakdown by race/ethnic group for the following baseline characteristics: age, gender, obesity class, medical comorbidities, distance to primary/tertiary care, socioeconomic status (zip code-based income quartile), zip code-based education, and rurality.

We visualized variability in use of weight loss medications regionally across the United States, using VA-defined geographic regions.

We then used separate generalized linear regression models with binary distributions to evaluate the association between use of the three categories of weight loss interventions (MOVE!, medication prescriptions, and bariatric surgery) and race/ethnicity. Multivariable logistic regression was performed to evaluate associations between weight loss interventions by race/ethnicity. Model covariate selection was performed with LASSO, which yielded inclusion of initial obesity class (overweight, Class I, Class II, and Class III), gender, age at diagnosis, race/ethnicity, rurality, education status, diabetes status, and Charlson Comorbidity Index (CCI) in the final adjusted models in SAS statistical software with diagnosis year as random effect. An additional multivariate logistic regression model added an interaction term for race/ethnicity and obesity class. Finally, we performed a sensitivity analysis limiting obesity medications to semaglutide only, and another excluding semaglutide, to evaluate the extent to which semaglutide was driving results.

To further examine the relationship between race/ethnicity and semaglutide use over time, we limited the existing cohort to only those patients with new diagnoses in the study period, defined as having no BMI measurements in the ten years prior to the study period. We then quantified the proportion of patients receiving at least one prescription for semaglutide in each calendar year in the study period, by race/ethnicity group.

This study was approved by the Stanford Institutional Review Board and the Veterans Affairs Research and Development Committee. Requirement for informed consent was waived.

## Results

### Obesity Prevalence and Demographics

Among Veterans with BMI ≥27 (n=2,423,070), 38.3% had Class 1 obesity (BMI 30-34.9), 18.1% had Class 2 obesity (BMI 35-39.9), and 9.8% had Class 3 obesity (BMI≥40). All race/ethnicity groups, apart from Asian/NHOPI, had a plurality of individuals with Class I obesity and successively smaller proportions with overweight and Class II and II obesity; however, Asian/NHOPI Veterans had the largest proportion falling in the Overweight category, with correspondingly smaller proportions of individuals with Class I, II or III obesity (**Figure 1**).

**Figure 1.**
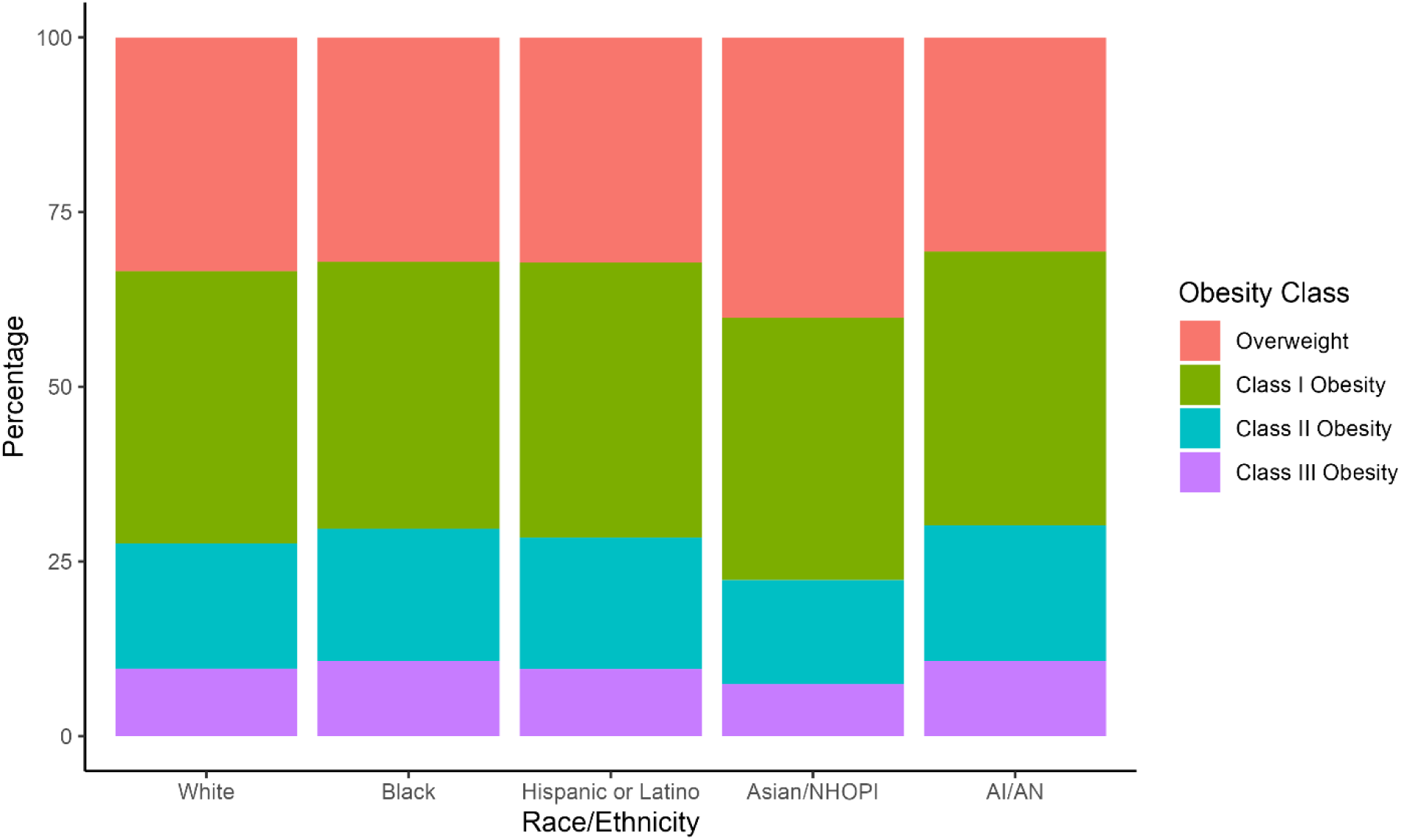
Obesity Class Distribution by Race/Ethnicity Distribution of individuals with overweight/obesity across overweight/obesity classes, by race/ethnicity.

Compared to all other groups, White patients tended to be older (**Table 1**). Women made up the smallest proportion of White patients (5.7%) and the largest proportion of Black patients (15.1%). Black and AI/AN patients had the highest proportions of severe (Class III) obesity at 10.8% in both groups, whereas Asian/NHOPI patients had the lowest (7.4%). White and Asian/NHOPI patients had larger proportions of individuals living in counties where <25% of inhabitants had a high school education or less (87.8 and 90.1%, respectively) compared to Black, Hispanic, and AI/AN patients (83.8, 79.5, and 83.1%, respectively). AI/AN patients had longer drive distances to primary and tertiary care than did other groups and larger proportions of rural or highly rural individuals. This group also had the highest rate of diabetes and the second-highest rate of chronic pulmonary disease.

**Table 1.** Baseline Patient Characteristics by Race/Ethnicity.

|  | Race/Ethnicity |  |  |  |  |  |  |
| --- | --- | --- | --- | --- | --- | --- | --- |
|  | All | White | Black | Hispanic or Latino | Asian/ NHOPI | AI/AN | P-value |
| **Total** | 2423070 | 1729905 | 493977 | 144543 | 37087 | 17558 |  |
| **Age at Diagnosis** |  |  |  |  |  |  | <.0001<sup>†</sup> |
| Mean (SD) | 63.6 (13.3) | 65.5 (13.1) | 59.2 (12.0) | 57.9 (14.4) | 58.8 (14.4) | 61.6 (13.3) |  |
| **Gender, n (%)** |  |  |  |  |  |  | <.0001<sup>‡</sup> |
| F | 189616<br>(7.8%) | 98249<br>(5.7%) | 74558<br>(15.1%) | 11582<br>(8.0%) | 3535<br>(9.5%) | 1692<br>(9.6%) |  |
| M | 2233454<br>(92.2%) | 1631656<br>(94.3%) | 419419<br>(84.9%) | 132961<br>(92.0%) | 33552<br>(90.5%) | 15866<br>(90.4%) |  |
| **Obesity class, n (%)** |  |  |  |  |  |  | <.0001<sup>‡</sup> |
| Overweight | 804022<br>(33.2%) | 578575<br>(33.4%) | 158642<br>(32.1%) | 46559<br>(32.2%) | 14866<br>(40.1%) | 5380<br>(30.6%) |  |
| Class I Obesity | 940827<br>(38.8%) | 674295<br>(39.0%) | 188806<br>(38.2%) | 56908<br>(39.4%) | 13939<br>(37.6%) | 6879<br>(39.2%) |  |
| Class II Obesity | 439763<br>(18.1%) | 310305<br>(17.9%) | 93350<br>(18.9%) | 27189<br>(18.8%) | 5514<br>(14.9%) | 3405<br>(19.4%) |  |
| Class III Obesity | 238458<br>(9.8%) | 166730<br>(9.6%) | 53179<br>(10.8%) | 13887<br>(9.6%) | 2768<br>(7.5%) | 1894<br>(10.8%) |  |
| **Comorbidities** |  |  |  |  |  |  |  |
| Charlson Comorbidity Index (CCI) |  |  |  |  |  |  | <.0001<sup>†</sup> |
| Mean (SD) | 1.7 (2.1) | 1.7 (2.1) | 1.7 (2.2) | 1.4 (2.0) | 1.4 (1.9) | 1.6 (2.1) |  |
| Median (Range) | 1.0 (0.0, 23.0) | 1.0 (0.0, 23.0) | 1.0 (0.0, 22.0) | 1.0 (0.0, 23.0) | 1.0 (0.0, 20.0) | 1.0 (0.0, 21.0) |  |
| Diabetes, n (%)* | 933119<br>(38.2%) | 657286<br>(37.7%) | 197288<br>(39.7%) | 56470<br>(38.8%) | 14727<br>(39.3%) | 7348<br>(41.6%) | <.0001<sup>‡</sup> |
| Cancer, n (%) | 207273<br>(8.5%) | 153641<br>(8.8%) | 42267<br>(8.5%) | 8175<br>(5.6%) | 2049<br>(5.5%) | 1141<br>(6.5%) | <.0001<sup>‡</sup> |
| Cerebrovascular Disease, n (%) | 161686<br>(6.6%) | 122332<br>(7.0%) | 29448<br>(5.9%) | 7024<br>(4.8%) | 1837<br>(4.9%) | 1045<br>(5.9%) | <.0001<sup>‡</sup> |
| Chronic Kidney Disease, n (%) | 246696<br>(10.1%) | 171402<br>(9.8%) | 59445<br>(12.0%) | 10880<br>(7.5%) | 3451<br>(9.2%) | 1518<br>(8.6%) | <.0001<sup>‡</sup> |
| Chronic Pulmonary Disease, n (%) | 429622<br>(17.6%) | 333500<br>(19.1%) | 70413<br>(14.2%) | 17544<br>(12.1%) | 5002<br>(13.4%) | 3163<br>(17.9%) | <.0001<sup>‡</sup> |
| Congestive Heart Failure, n (%) | 186841<br>(7.7%) | 139442<br>(8.0%) | 36894<br>(7.4%) | 7209<br>(5.0%) | 2051<br>(5.5%) | 1245<br>(7.0%) | <.0001<sup>‡</sup> |
| Hyperlipidemia, n (%) | 1566355<br>(64.6%) | 1169855<br>(67.6%) | 276636<br>(56.0%) | 86202<br>(59.6%) | 22836<br>(61.6%) | 10826<br>(61.7%) | <.0001<sup>‡</sup> |
| Hypertension, n (%) | 1814003<br>(74.9%) | 1292200<br>(74.7%) | 386380<br>(78.2%) | 95937<br>(66.4%) | 26792<br>(72.2%) | 12694<br>(72.3%) | <.0001<sup>‡</sup> |
| Metastatic Cancer, n (%) | 16718<br>(0.7%) | 12485<br>(0.7%) | 3240<br>(0.7%) | 689<br>(0.5%) | 191<br>(0.5%) | 113<br>(0.6%) | <.0001<sup>‡</sup> |
| Peripheral Vascular Disease, n (%) | 201975<br>(8.3%) | 160963<br>(9.2%) | 30581<br>(6.2%) | 7422<br>(5.1%) | 1832<br>(4.9%) | 1177<br>(6.7%) | <.0001<sup>‡</sup> |
| **Drive Distance to Primary Care** |  |  |  |  |  |  | <.0001<sup>†</sup> |
| Mean (SD) | 16.3 (15.9) | 17.7 (16.7) | 11.9 (10.8) | 13.6 (15.9) | 12.6 (12.8) | 23.7 (28.8) |  |
| **Drive Distance to** |  |  |  |  |  |  | <.0001<sup>†</sup> |
| **Tertiary Care** |  |  |  |  |  |  |  |
| Mean (SD) | 94.1 (81.5) | 101.4 (84.0) | 69.3 (61.1) | 91.4 (90.5) | 81.9 (79.7) | 129.7 (110.1) |  |
| **Education Level, n %** |  |  |  |  |  |  | <.0001<sup>‡</sup> |
| <25% of HS or less | 2066286<br>(86.5%) | 1499415<br>(87.8%) | 406438<br>(83.8%) | 113136<br>(79.5%) | 32977<br>(90.1%) | 14320<br>(83.0%) |  |
| >=25% of HS or less | 323005<br>(13.5%) | 208571<br>(12.2%) | 78646<br>(16.2%) | 29234<br>(20.5%) | 3623<br>(9.9%) | 2931<br>(17.0%) |  |
| **Income, % (n)** |  |  |  |  |  |  | <.0001<sup>‡</sup> |
| <=25% 75K or less | 1862009<br>(78.0%) | 1323473<br>(77.5%) | 394083<br>(81.3%) | 108057<br>(75.9%) | 21964<br>(60.0%) | 14432<br>(83.8%) |  |
| >25% 75K or less | 525676<br>(22.0%) | 383542<br>(22.5%) | 90499<br>(18.7%) | 34221<br>(24.1%) | 14621<br>(40.0%) | 2793<br>(16.2%) |  |
| **Rurality, n (%)** |  |  |  |  |  |  | <.0001<sup>†</sup> |
| Highly Rural | 92663<br>(4.0%) | 85004<br>(5.1%) | 4153<br>(0.9%) | 1677<br>(1.2%) | 442<br>(1.4%) | 1387<br>(8.3%) |  |
| Rural | 740890<br>(31.9%) | 623582<br>(37.6%) | 80844<br>(16.9%) | 24280<br>(17.4%) | 5320<br>(16.5%) | 6864<br>(41.2%) |  |
| Urban | 1491306<br>(64.1%) | 949010<br>(57.3%) | 393773<br>(82.2%) | 113641<br>(81.4%) | 26489<br>(82.1%) | 8393<br>(50.4%) |  |

\*In contrast to other comorbidities, which are included for the pre-study period only, diabetes was included if patient had this diagnosis before or during the study period; <sup>†</sup> Kruskal-Wallis p-value; <sup>‡</sup> Chi-Square p-value

Patients with Class III obesity (BMI ≥40) were younger and had more medical comorbidities (higher CCI) than those with lower BMI. Women Veterans, those who were unmarried, and those living in rural locations were also disproportionately represented in the group with Class III obesity (**Table S3**). AI/AN and Black Veterans’ BMI categories also skewed toward Class III obesity; this was most significant for Black patients, who comprised 20.4% of the overall cohort and 22.3% of those with Class III obesity, whereas AI/AN individuals comprised 0.8% of the group with Class III obesity and 0.7% of the overall cohort. Asian/NHOPI Veterans, conversely, comprised 1.5% of those with Class III obesity but 1.2% of the overall cohort; Hispanic/Latino and White individuals similarly had slightly lower representation among those with Class III obesity versus in the overall cohort. More than half (50.9%) of those with Class III obesity had a diagnosis of diabetes, compared to 31.7% of those with overweight, 37.9% of those with Class I obesity, and 44.9% of those with Class II obesity.

### Use of Interventions

Rates of use of all three types of interventions were low (**Table 2, Table S4**). Across the cohort, 7.7% participated in MOVE!, 7.9% received weight loss medications, and 0.1% underwent bariatric surgery. Semaglutide was the most frequent weight loss medication used, with 96% (n=184,690) medication users filling a prescription for semaglutide at least once during the study time period (**Table S5**). Use of medications varied within a relatively narrow range by region (**Figure S1**).

**Table 2.** Intervention Use by Race/Ethnicity.

|  | **Race/Ethnicity** |  |  |  |  |  |  |
| --- | --- | --- | --- | --- | --- | --- | --- |
|  | All | White | Black | Hispanic or Latino | Asian/NHOPI | AI/AN | P-value |
| **Total** | 2423070 | 1729905 | 493977 | 144543 | 37087 | 17558 |  |
| **Interventions** |  |  |  |  |  |  |  |
| MOVE! | 186124<br>(7.7%) | 114575<br>(6.6%) | 52353<br>(10.6%) | 14508<br>(10.0%) | 3239<br>(8.7%) | 1449<br>(8.3%) | <0.01 |
| Medications | 192487<br>(7.5%) | 135183<br>(7.8%) | 39556<br>(8.0%) | 12961<br>(9.0%) | 3329<br>(9.0%) | 1458<br>(8.3%) | <0.01 |
| Bariatric Surgery | 2806<br>(0.1%) | 1808<br>(0.1%) | 711<br>(0.1%) | 233<br>(0.2%) | 35<br>(0.1%) | 19<br>(0.1%) | <0.01 |
| **Combinations** |  |  |  |  |  |  |  |
| MOVE! and meds | 34851<br>(1.4%) | 23030<br>(1.3%) | 8357<br>(1.6%) | 2594<br>(1.7%) | 580<br>(1.5%) | 290<br>(1.6%) | <0.01 |
| MOVE! and surgery | 1034<br>(0.04%) | 701<br>(0.03%) | 231<br>(0.03%) | 91<br>(0.03%) | 8<br>(0.03%) | 3<br>(0.0%) | <0.01 |
| Meds and surgery | 894<br>(0.04%) | 558<br>(0.02%) | 246<br>(0.03%) | 76<br>(0.05%) | 7<br>(0.11%) | 7<br>(0.02%) | <0.01 |
| MOVE!, meds, and surgery | 344<br>(0.01%) | 230<br>(0.01%) | 81<br>(0.02%) | 28<br>(0.02%) | 2<br>(0.01%) | 3<br>(0.01%) | <0.01 |
| Medications only | 157086<br>(6.5%) | 111825<br>(6.5%) | 31034<br>(6.3%) | 10319<br>(7.1%) | 2744<br>(7.4%) | 1164<br>(6.6%) | <0.01 |
| Bariatric Surgery only | 1222<br>(0.05%) | 779<br>(0.05%) | 315<br>(0.06%) | 94<br>(0.07%) | 22<br>(0.06%) | 12<br>(0.07%) | <0.01 |
| MOVE! only | 150583<br>(6.2%) | 91074<br>(5.3%) | 43846<br>(8.9%) | 118591<br>(8.2%) | 2653<br>(7.2%) | 1159<br>(6.6%) | <0.01 |

Use of MOVE! ranged from 6.6% for White patients to 10.6% for Black patients (**Table 2**). Medication use also varied but within a smaller range, with the lowest use among White patients at 7.8% and highest among Hispanic/Latino and Asian/NHOPI patients at 9.0%. Bariatric surgery use ranged between 0.1-0.2% across all race/ethnicity groups. Combinations of interventions were relatively uncommon; for example, just 1.4% of the cohort used both MOVE! and medications within the study period.

Adjusted odds of accessing a given intervention increased with obesity class for all three interventions (**Table S6**), with the most dramatic gradient for bariatric surgery: compared to those with overweight, Veterans with class II obesity had adjusted odds of undergoing bariatric surgery of 5.87, and those with class III obesity adjusted odds of 27.4. Compared to men, adjusted odds of accessing all three interventions were significantly higher for women: for MOVE!, odds ratio (OR) 1.92 (95% CI 1.89, 1.95); for medications, OR 1.77 (95% CI 1.75, 1.81); for bariatric surgery, OR 3.14 (95% CI 2.88, 3.41).

We found significant differences by race/ethnicity for all three types of interventions that persisted after adjustment (**Figure 2**). Compared to White patients, patients belonging to racial/ethnic minority groups other than AI/AN were more likely to participate in MOVE! (Asian/NHOPI Veterans, 1.12 [95% CI 1.06-1.19]; Black Veterans, OR 1.24 [95% confidence interval [CI] 1.22-1.26]; Hispanic/Latino Veterans, OR 1.17 [95% CI 1.14-1.20]). Patients in all non-White groups were less likely to receive weight loss medications (AI/AN Veterans, OR 0.84 [95% CI 0.77-0.92]; Asian/NHOPI Veterans: OR 0.94 [95% CI 0.89-0.999]; Black Veterans, OR 0.75 [95% CI 0.74-0.76]; Hispanic/Latino Veterans, adj OR 0.94 [95% CI 0.91-0.97]). The adjusted odds of receiving bariatric surgery for Black patients were significantly lower than for White patients (OR 0.79 [95% CI 0.69- 0.89]). No differences in receipt of bariatric surgery were seen for other racial/ethnic groups.

**Figure 2.**
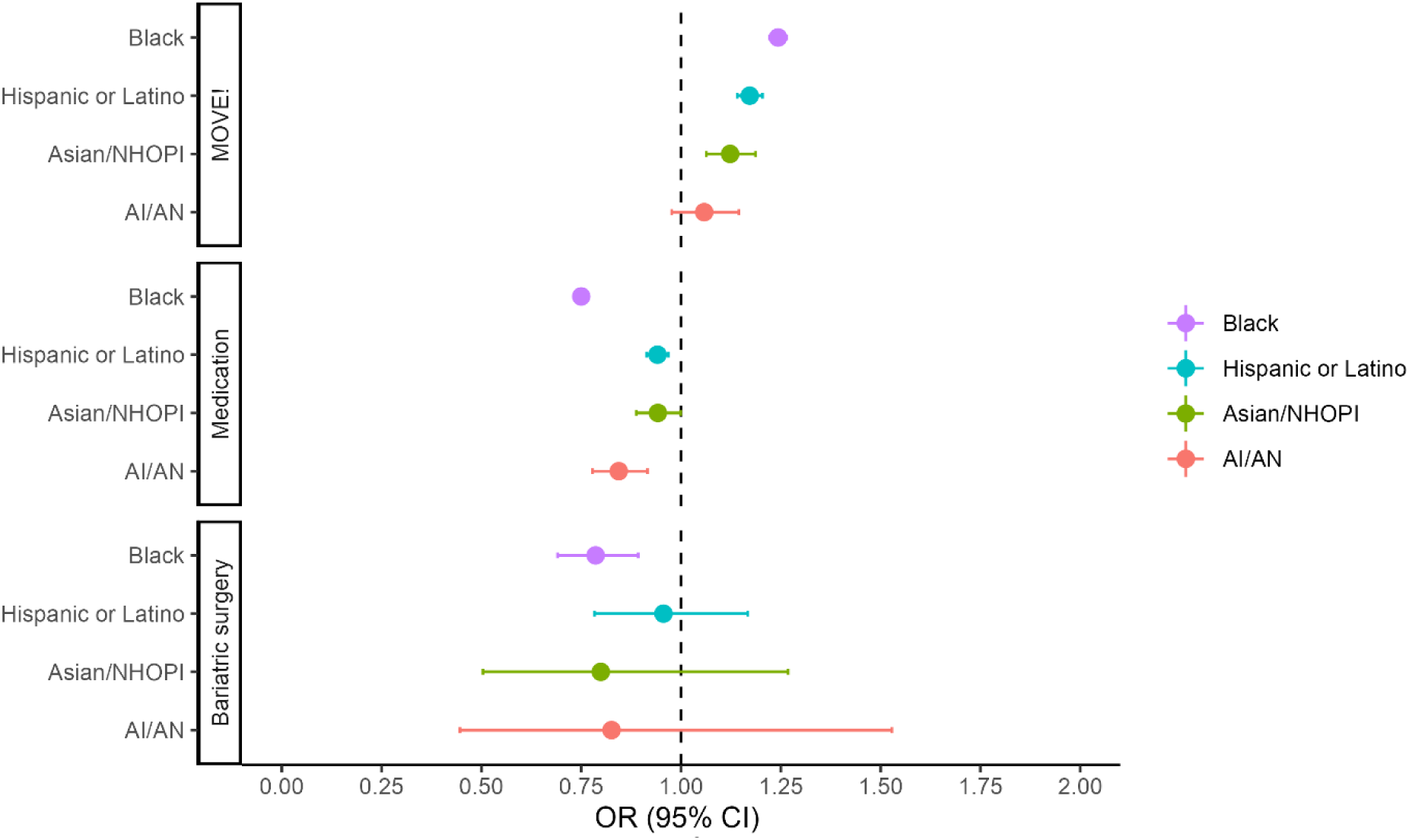
Adjusted Odds of Receiving Obesity Treatments by Race/Ethnicity AI = American Indian; AN = Alaska Native; NHOPI = Native Hawaiian or Pacific Islander. Reference race/ethnicity group is White.

In models containing an interaction term for race/ethnicity x obesity class (**Table S7**), differences between minoritized and White patients grew smaller with higher obesity class for MOVE! use for Asian/NHOPI, Black, and Hispanic/Latino patients. For medications and bariatric surgery, however, there were no significant differences in the severity of inequities in use between minoritized and White patients across obesity classes.

### Sensitivity Analyses

In a statistical model including semaglutide as the only medication type (**Table S8**) and adjusting for similar covariates, results for race/ethnicity were largely unchanged, except that the AI/AN and Asian/NHOPI race/ethnicity group findings were no longer significant (AI/AN patients, OR 0.82 [95% CI 0.63-1.07; Asian/NHOPI patients, OR 0.96 [95% CI 0.80-1.14]; Black patients, OR 0.63 [95% CI 0.60-0.68); Hispanic/Latino patients, OR 0.81 (95% CI 0.74-0.90]). Carrying a diagnosis of diabetes was associated with higher use of semaglutide, with an odds ratio of 35.2 (95% CI 32.5-38.0) compared to those without diabetes. In the converse model excluding semaglutide (**Table S8**), there was a stronger effect of obesity class; e.g., the odds ratio for use of medications for those with Class III obesity compared to those with overweight was 14.3 (95% CI 13.3-15.3) versus just 3.32 in the semaglutide-users-only model (95% CI 3.01-3.56). Adjusted odds ratios for minority race/ethnicity groups were still significantly lower than for the White comparison group, but the effect was attenuated for the Black and Hispanic/Latino groups compared to the primary or semaglutide-users-only models (Black patients, OR 0.94 [95% CI 0.89-0.97]; Hispanic/Latino patients, OR 0.88 [95% CI 0.81-0.95]).

### Use of Semaglutide over Time

In our analysis of semaglutide use by race/ethnicity group over time, overall use of semaglutide increased significantly over the study period (**Figure 3**, **Table S9**). However, only Black patients had semaglutide use rates that were significantly lower than those of White patients in several study years.

**Figure 3.**
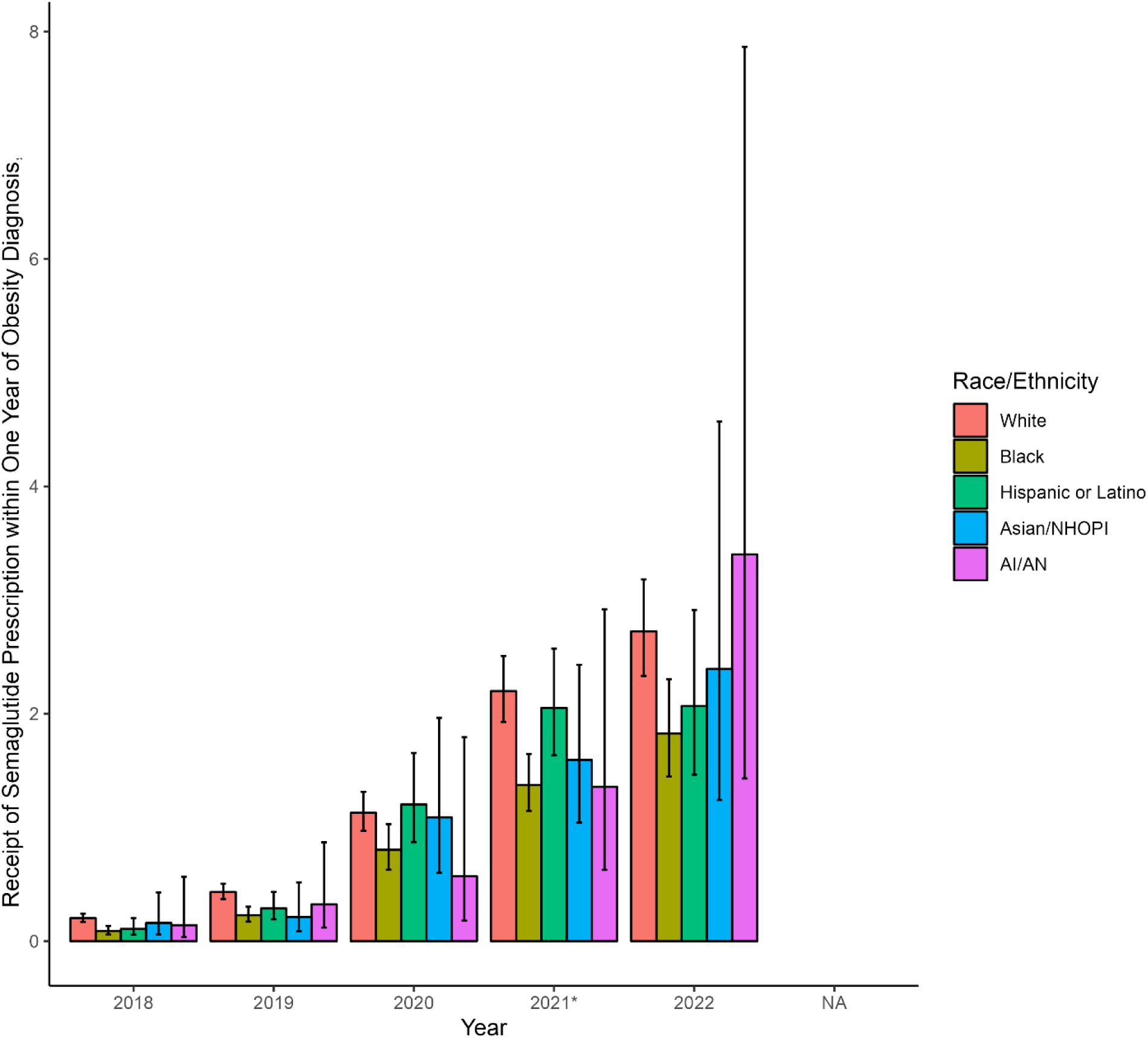
Semaglutide Use by Race/Ethnicity Over Time Distribution of individuals with overweightPercentage of individuals with overweight/obesity receiving semaglutide within one year of diagnosis. *Semaglutide received FDA labeling for chronic weight management in July 2021.

## Discussion

In this analysis of contemporary obesity treatments among over 2.4 million eligible VA patients, we have identified low uptake and significant differences by race/ethnicity in use of lifestyle change counseling, weight loss medications, and bariatric surgery. These inequities persist across the spectrum of severity of obesity. The VA system has been a leader in efforts to reduce inequities with some success (32), and these findings offer another challenge to those efforts.

The proportion of eligible Veterans found to be using the MOVE! program and bariatric surgery are closely in line with past estimates (13,17), with at most a slight increase in bariatric surgery in recent years. Conversely, use of weight loss medications among Veterans, though still low overall, has increased significantly in recent years, from estimates in the range of 1-2% of eligible Veterans in studies examining use in the early-mid 2010s (14,19) to almost 8% in the present study. This trend is expected against the backdrop development of novel and effective weight loss medications (i.e., GLP-1 receptor agonists) emerging over this time period. Despite FDA labeling of semaglutide for weight loss that was limited to the latter half of our study period, our sensitivity analyses suggest that semaglutide use largely drove our overall medication-related results.

Notwithstanding this modest increase in medication use over time, it is sobering that the overall use of guideline-recommended (8–11) and other evidence-based (33,34) interventions for obesity remains at such low levels despite the high morbidity and cost associated with this condition. These low rates of obesity intervention uptake persist even in the VA population who have universal access to free or low-cost healthcare. Given well-publicized gaps in insurance coverage for novel—or indeed, any— weight loss agents (35–37), it is likely that access suffers more in the general population (26).

Our results reveal higher adjusted odds of using MOVE! and lower odds of using medications for several minoritized patient groups, as well as lower odds of undergoing bariatric surgery for Black patients. While MOVE! and similar lifestyle counseling interventions are evidence-based avenues to weight loss, medications and surgery produce substantially more weight loss on average (12,37). Thus, this set of results indicating lower access to more intensive, more effective treatments among minoritized patients is troubling, even if access to MOVE! may be better than it is for White patients. Unfortunately, these findings are consistent with a growing body of work demonstrating worse access to evidence-based therapies for cardiometabolic disease among, in particular, Black and Hispanic individuals across a host of conditions in and outside VA (38–41). Though these disparities are often attributed to differences in insurance coverage or socioeconomic status, their persistence in an integrated healthcare system with minimal cost-sharing, such as the VA, suggests more complex barriers to care. Mechanisms of systemic racism that may contribute to these patterns must be further investigated and dismantled. Analogous to the overall low uptake of obesity interventions discussed above, these inequities are likely to be worse in systems without such mechanisms to level the financial playing field (42). Indeed, disparities have been demonstrated among patients taking GLP-1 receptor agonists for diabetes (38,43).

Another problematic aspect of our results is the finding that individuals in the “Asian/NHOPI” race/ethnicity category had lower use of medications than White patients. These results raise concern that patients of Asian backgrounds may not be accessing treatment at lower BMIs as would be recommended based on this population’s disproportionately high cardiovascular risk (44–46). Further granular research on this high risk population is warranted.

Both the low overall use of obesity interventions and inequities therein are problematic and require intervention. One strategy is to leverage pandemic-era developments in virtual care overall (47) and within obesity medicine (48,49). Past work suggests that provision of obesity interventions varies significantly across VA sites (13,16). Capitalizing on the prevalence of virtual care use in VA and the emergence of hub-and-spoke virtual clinics could extend the reach of these services from VA centers with stronger obesity medicine programs to relatively underserved facilities, with the caveat that hub sites will need to be adequately resourced to be able to serve these larger catchment areas. Second, offering other categories of interventions to individuals already engaged in one type of intervention (e.g., adding medications for those in the MOVE! program) would likely increase the proportion of eligible patients accessing a given type of intervention, given we found low use of combinations of interventions. Third, as regulations surrounding provision of obesity interventions change with the evolving pharmacological and procedural landscape, equity concerns must be prioritized. While our study period concluded at the end of 2022, the landscape of weight loss medication availability has been dynamic in the months and years immediately preceding the writing of this manuscript: semaglutide’s popularity produced global supply shortages limiting new starts of the medication throughout much of 2023, and tirzepatide received FDA labeling for weight loss in November 2023 (2). Given the inequities our study demonstrates, it is incumbent upon clinicians and policymakers to include equity concerns in any approach to prescribing these novel, and effectively scarce, agents.

### Limitations

Many obesity medications are used for multiple indications. As our data did not include detailed indication information from pharmacy dispensing records, we cannot precisely capture when medications are being used for obesity versus for another indication. In particular, we cannot differentiate whether the primary indication for semaglutide use was diabetes versus obesity, though we adjusted for diabetes status to mitigate this concern. Similarly, while the Contrave combination bupropion/naltrexone medication is labeled only for obesity, it is less clear whether the indication for these medications is obesity when they are used as their individual components of bupropion and naltrexone. There may also be additional medications used on- or off-label for obesity that are not captured in our analysis–most notably liraglutide, which we excluded since this medication is explicitly not preferred for weight management within VA. Our method of defining the interventions excludes those who used MOVE! or surgery within the five years prior to the study period in order to capture more recent trends in use of these interventions; this method may result in an underestimate of the total number of Veterans with overweight or obesity accessing these interventions, and the true use of MOVE! and/or bariatric surgery may be higher than our results suggest. Reassuringly, our estimates of MOVE! and bariatric surgery use are similar to others’ within VA (13,17). Our analysis does not extend to more novel surgical or endoscopic interventions, however given data suggesting late adoption of novel therapies among minoritized individuals (21,50), we expect this information would only exacerbate the severity of inequities identified rather than change the findings. Finally, we cannot determine the extent to which whether the differences we see between race/ethnicity groups are attributable to differential treatment preferences by race/ethnicity, or other mechanisms.

## Conclusion

Inequities by race/ethnicity exist across the spectrum of obesity interventions in this population of 2.4 million Veterans eligible for such services, with minoritized individuals less likely to receive evidence-based approaches to treatment despite the lack of insurance barriers in this population—particularly novel anti-obesity medications. As the field of obesity medicine evolves at an unprecedented pace, consideration of the mechanisms for equitable implementation of novel therapies will be critical to close widening gaps in care.

### Sources of Funding

CMY is funded by a VA Career Development Award from the U.S. Department of Veterans Affairs, Health Services Research & Development Service of the VA Office of Research and Development.

## Data Availability

Patient-level data cannot be made available by the authors to other researchers per VA policy. However, all data used in the analyses are available to VA researchers through the VA Informatics and Computing Infrastructure.

## References

1. Office of the Commissioner of the FDA. FDA. FDA; 2021 [cited 2024 Jan 12]. FDA Approves New Drug Treatment for Chronic Weight Management, First Since 2014. Available from: https://www.fda.gov/news-events/press-announcements/fda-approves-new-drug-treatment-chronic-weight-management-first-2014

2. Office of the Commissioner of the FDA. FDA. 2023 [cited 2024 Jan 12]. FDA Approves New Medication for Chronic Weight Management. Available from: https://www.fda.gov/news-events/press-announcements/fda-approves-new-medication-chronic-weight-management

3. Stierman B, Afful J, Carroll MD, Chen TC, Davy O, Fink S, et al. National Health and Nutrition Examination Survey 2017–March 2020 Prepandemic Data Files Development of Files and Prevalence Estimates for Selected Health Outcomes. Natl Health Stat Rep. 2021 Jun 14;(158).

4. Office of the Surgeon General (US), Office of Disease Prevention and Health Promotion (US), Centers for Disease Control and Prevention (US), National Institutes of Health (US). Section 1: Overweight and Obesity as Public Health Problems in America. In: The Surgeon General’s Call To Action To Prevent and Decrease Overweight and Obesity [Internet]. Office of the Surgeon General (US); 2001 [cited 2024 Jan 22]. Available from: https://www.ncbi.nlm.nih.gov/books/NBK44210/

5. The GBD 2015 Obesity Collaborators. Health Effects of Overweight and Obesity in 195 Countries over 25 Years. N Engl J Med. 2017 Jul 6;377(1):13–27.

6. Breland JY, Phibbs CS, Hoggatt KJ, Washington DL, Lee J, Haskell S, et al. The Obesity Epidemic in the Veterans Health Administration: Prevalence Among Key Populations of Women and Men Veterans. J Gen Intern Med. 2017 Apr 1;32(1):11–7.

7. Ward ZJ, Bleich SN, Cradock AL, Barrett JL, Giles CM, Flax C, et al. Projected U.S. State-Level Prevalence of Adult Obesity and Severe Obesity. N Engl J Med. 2019 Dec 19;381(25):2440–50.

8. Jensen MD, Ryan DH, Apovian CM, Ard JD, Comuzzie AG, Donato KA, et al. 2013 AHA/ACC/TOS Guideline for the Management of Overweight and Obesity in Adults. Circulation. 2014 Jun 24;129(25 Suppl 2):S102–38.

9. Apovian CM, Aronne LJ, Bessesen DH, McDonnell ME, Murad MH, Pagotto U, et al. Pharmacological Management of Obesity: An Endocrine Society Clinical Practice Guideline. J Clin Endocrinol Metab. 2015 Feb;100(2):342–62.

10. Garvey WT, Mechanick JI, Brett EM, Garber AJ, Hurley DL, Jastreboff AM, et al. American Association of Clinical Endocrinologists and American College of Endocrinology Comprehensive Clinical Practice Guidelines For Medical Care of Patients with Obesity. Endocr Pract. 2016 Jul 1;22:1–203.

11. Eisenberg D, Shikora SA, Aarts E, Aminian A, Angrisani L, Cohen RV, et al. 2022 American Society of Metabolic and Bariatric Surgery (ASMBS) and International Federation for the Surgery of Obesity and Metabolic Disorders (IFSO) Indications for Metabolic and Bariatric Surgery. Obes Surg. 2023 Jan 1;33(1):3– 14.

12. Elmaleh-Sachs A, Schwartz JL, Bramante CT, Nicklas JM, Gudzune KA, Jay M. Obesity Management in Adults: A Review. JAMA. 2023 Nov 28;330(20):2000–15.

13. Maciejewski ML, Arterburn DE, Berkowitz TSZ, Weidenbacher HJ, Liu CF, Olsen MK, et al. Geographic Variation in Obesity, Behavioral Treatment, and Bariatric Surgery for Veterans. Obesity. 2019;27(1):161–5.

14. Semla TP, Ruser C, Good CB, Yanovski SZ, Ames D, Copeland LA, et al. Pharmacotherapy for Weight Management in the VHA. J Gen Intern Med. 2017 Apr 1;32(1):70–3.

15. Claridy MD, Czepiel KS, Bajaj SS, Stanford FC. Treatment of Obesity: Pharmacotherapy Trends of Office-Based Visits in the United States From 2011 to 2016. Mayo Clin Proc. 2021 Dec 1;96(12):2991–3000.

16. Gunnar W. Bariatric Surgery Provided by the Veterans Health Administration: Current State and a Look to the Future. J Gen Intern Med. 2017 Apr 1;32(1):4–5.

17. Maciejewski ML, Shepherd-Banigan M, Raffa SD, Weidenbacher HJ. Systematic Review of Behavioral Weight Management Program MOVE! for Veterans. Am J Prev Med. 2018 May 1;54(5):704–14.

18. Robinson KM, Vander Weg M, Laroche HH, Carrel M, Wachsmuth J, Kazembe K, et al. Obesity treatment initiation, retention, and outcomes in the Veterans Affairs MOVE! Program among rural and urban veterans. Obes Sci Pract [Internet]. 2022 Jun 8 [cited 2022 Nov 23]; Available from: http://onlinelibrary.wiley.com/doi/abs/10.1002/osp4.622

19. Thomas DD, Waring ME, Ameli O, Reisman JI, Vimalananda VG. Patient Characteristics Associated with Receipt of Prescription Weight-Management Medications Among Veterans Participating in MOVE! Obesity. 2019;27(7):1168–76.

20. Dearing JW, Cox JG. Diffusion Of Innovations Theory, Principles, And Practice. Health Aff (Millwood). 2018 Feb;37(2):183–90.

21. Yong CM, Jaluba K, Batchelor W, Gummipundi S, Asch SM, Heidenreich P. Temporal trends in transcatheter aortic valve replacement use and outcomes by race, ethnicity, and sex. Catheter Cardiovasc Interv. 2022;99(7):2092–100.

22. Bhogal SK, Reddigan JI, Rotstein OD, Cohen A, Glockler D, Tricco AC, et al. Inequity to the Utilization of Bariatric Surgery: a Systematic Review and Meta-Analysis. Obes Surg. 2015 May 1;25(5):888–99.

23. Funk LM, Alagoz E, Murtha JA, Breuer CR, Pati B, Eierman L, et al. Socioeconomic disparities and bariatric surgery outcomes: A qualitative analysis. Am J Surg [Internet]. 2022 Sep 27 [cited 2022 Nov 23]; Available from: https://www.sciencedirect.com/science/article/pii/S0002961022006195

24. Flum DR, Khan TV, Dellinger EP. Toward the Rational and Equitable Use of Bariatric Surgery. JAMA. 2007 Sep 26;298(12):1442.

25. Martin M, Beekley A, Kjorstad R, Sebesta J. Socioeconomic disparities in eligibility and access to bariatric surgery: a national population-based analysis. Surg Obes Relat Dis. 2010 Jan 1;6(1):8–15.

26. Waldrop SW, Johnson VR, Stanford FC. Inequalities in the provision of GLP-1 receptor agonists for the treatment of obesity. Nat Med. 2024 Jan 3;1–4.

27. Veterans Health Administration. About VHA [Internet]. [cited 2024 Feb 1]. Available from: https://www.va.gov/health/aboutvha.asp

28. CDC. Centers for Disease Control and Prevention. 2022 [cited 2024 Jan 16]. All About Adult BMI. Available from: https://www.cdc.gov/healthyweight/assessing/bmi/adult_bmi/index.html

29. Maciejewski ML, Yancy WS, Olsen M, Weidenbacher HJ, Abbott D, Weinberger M, et al. Demand for Weight Loss Counseling After Copayment Elimination. Prev Chronic Dis. 2013 Apr 4;10:E49.

30. Kinsinger LS, Jones KR, Kahwati L, Harvey R, Burdick M, Zele V, et al. Design and Dissemination of the MOVE! Weight-Management Program for Veterans. Prev Chronic Dis. 2009 Jun 15;6(3):A98.

31. U.S. Department of Veterans Affairs. MOVE! Weight Management Program [Internet]. [cited 2024 Jan 12]. Available from: https://www.move.va.gov/

32. Ibrahim SA, Egede LE, Uchendu US, Fine MJ. The Struggle for Health Equity: The Sustained Effort by the VA Healthcare System. Am J Public Health. 2014 Sep;104(S4):S514–6.

33. Wilding JPH, Batterham RL, Calanna S, Davies M, Van Gaal LF, Lingvay I, et al. Once-Weekly Semaglutide in Adults with Overweight or Obesity. N Engl J Med. 2021 Mar 18;384(11):989–1002.

34. Jastreboff AM, Aronne LJ, Ahmad NN, Wharton S, Connery L, Alves B, et al. Tirzepatide Once Weekly for the Treatment of Obesity. N Engl J Med. 2022 Jul 21;387(3):205–16.

35. Gomez G, Stanford FC. US health policy and prescription drug coverage of FDA-approved medications for the treatment of obesity. Int J Obes. 2018 Mar;42(3):495–500.

36. Baum C, Andino K, Wittbrodt E, Stewart S, Szymanski K, Turpin R. The Challenges and Opportunities Associated with Reimbursement for Obesity Pharmacotherapy in the USA. PharmacoEconomics. 2015 Jul 1;33(7):643–53.

37. Bessesen DH, Van Gaal LF. Progress and challenges in anti-obesity pharmacotherapy. Lancet Diabetes Endocrinol. 2018 Mar 1;6(3):237–48.

38. Eberly LA, Yang L, Essien UR, Eneanya ND, Julien HM, Luo J, et al. Racial, Ethnic, and Socioeconomic Inequities in Glucagon-Like Peptide-1 Receptor Agonist Use Among Patients With Diabetes in the US. JAMA Health Forum. 2021 Dec 17;2(12):e214182.

39. Essien UR, Kim N, Hausmann LRM, Mor MK, Good CB, Magnani JW, et al. Disparities in Anticoagulant Therapy Initiation for Incident Atrial Fibrillation by Race/Ethnicity Among Patients in the Veterans Health Administration System. JAMA Netw Open. 2021 Jul 28;4(7):e2114234.

40. Essien UR, Holmes DN, Jackson LR, Fonarow GC, Mahaffey KW, Reiffel JA, et al. Association of Race/Ethnicity With Oral Anticoagulant Use in Patients With Atrial Fibrillation. JAMA Cardiol. 2018 Dec;3(12):1174–82.

41. Nathan Ashwin S., Raman Swathi, Yang Nancy, Painter Ian, Khatana Sameed Ahmed M., Dayoub Elias J., et al. Association Between 90-Minute Door-to-Balloon Time, Selective Exclusion of Myocardial Infarction Cases, and Access Site Choice. Circ Cardiovasc Interv. 2020 Sep 1;13(9):e009179.

42. Lu Y, Liu Y, Krumholz HM. Racial and Ethnic Disparities in Financial Barriers Among Overweight and Obese Adults Eligible for Semaglutide in the United States. J Am Heart Assoc. 2022 Oct 4;11(19):e025545.

43. Elhussein A, Anderson A, Bancks MP, Coday M, Knowler WC, Peters A, et al. Racial/ethnic and socioeconomic disparities in the use of newer diabetes medications in the Look AHEAD study. Lancet Reg Health - Am. 2022 Feb 1;6:100111.

44. Volgman AS, Palaniappan LS, Aggarwal NT, Gupta M, Khandelwal A, Krishnan AV, et al. Atherosclerotic Cardiovascular Disease in South Asians in the United States: Epidemiology, Risk Factors, and Treatments: A Scientific Statement From the American Heart Association. Circulation. 2018 Jul 3;138(1):e1–34.

45. Jih J, Mukherjea A, Vittinghoff E, Nguyen TT, Tsoh JY, Fukuoka Y, et al. Using appropriate body mass index cut points for overweight and obesity among Asian Americans. Prev Med. 2014 Aug;65:1–6.

46. Palaniappan LP, Wong EC, Shin JJ, Fortmann SP, Lauderdale DS. Asian Americans Have Greater Prevalence of Metabolic Syndrome Despite Lower Body Mass Index. Int J Obes 2005. 2011 Mar;35(3):393– 400.

47. Ferguson JM, Jacobs J, Yefimova M, Greene L, Heyworth L, Zulman DM. Virtual care expansion in the Veterans Health Administration during the COVID-19 pandemic: clinical services and patient characteristics associated with utilization. J Am Med Inform Assoc. 2020 Oct 30;ocaa284.

48. Kahan S, Look M, Fitch A. The benefit of telemedicine in obesity care. Obesity. 2022;30(3):577–86.

49. Lohnberg JA, Salcido L, Frayne S, Mahtani N, Bates C, Hauser ME, et al. Rapid conversion to virtual obesity care in COVID-19: Impact on patient care, interdisciplinary collaboration, and training. Obes Sci Pract. 2022;8(1):131–6.

50. Arozullah AM, Ferreira MR, Bennett RL, Gilman S, Henderson WG, Daley J, et al. Racial variation in the use of laparoscopic cholecystectomy in the Department of Veterans Affairs Medical System. J Am Coll Surg. 1999 Jun 1;188(6):604–22.

